# Impact of public health interventions on the COVID-19 epidemic: a stochastic model based on data from an African island

**DOI:** 10.1101/2020.06.22.20134130

**Authors:** D. C. Nuckchady

## Abstract

A stochastic model was created to simulate the impact of various healthcare measures on the COVID-19 epidemic. Travel restrictions and point of entry or exit screening help to delay the onset of the outbreak by a few weeks. Population surveillance is critical to detect the start of community transmission early and to avoid a surge in cases. Contact reduction and contact tracing are key interventions that can help to control the outbreak. To promptly curb the number of new cases, countries should diagnose patients using a highly sensitive test.

## Introduction

The effects of various public health strategies to halt the progression of Severe Acute Respiratory Syndrome Coronavirus 2 (SARS-CoV-2) are uncertain. This article discusses a simple mathematical and epidemiological model that simulates the transmission of SARS-CoV-2 in Mauritius. The outcomes of various testing strategies, contact tracing, point of entry screening, effective contact reduction and population-based surveillance are estimated using this model.

## Methods

Stochastic modeling is utilized in this article instead of a deterministic one since stochastic models are known to fail less often (1). Initially, a population of fixed size is assumed to be entirely susceptible to SARS-CoV-2. Infected people can enter the country at a pre-determined rate. Symptomatic patients visit a healthcare center and get tested. Those patients who are tested positive are isolated and started on treatment. If the test is negative, a repeat test is done – each patient can undergo a maximum of 2 tests. The threshold for testing each patient varies depending on the severity of his / her symptoms. Patients who die or who become immune to the disease are removed from the set of susceptible persons; however, a proportion of patients will not mount a proper immune response and will remain susceptible to the disease according to the model.

Several measures can be undertaken by the country to reduce the burden of SARS-CoV-2. These are travel restrictions, point of entry screening using questionnaires and non-contact thermometers, increasing the number of tests being done in the population, iterative contact tracing, quarantining all contacts and persons who fail the screening test, and putting in place strategies to effectively reduce contacts. The latter is not limited to community lockdown, broad confinement and universal curfew; any techniques that may be effective in reducing transmission of the disease during contacts can be considered e.g. unrestricted wearing of masks, prevalent hand hygiene, social distancing and proper cough etiquette. In most countries, such measures are intensified after the first few cases of COVID-19 have been diagnosed.

Figure 1 describes the schema used in the model. Patients who are isolated or in quarantine can still transmit the disease but at a much lower rate. The values of the variables used in the standard model are shown in table 1. The justification for employing these numbers is provided in section A of the accompanying appendix. Additional assumptions that were made in the model are specified in section B while the mathematical details are described in section C. The values were fitted to give a mean basic reproductive number R_0_ of 2.8 (mean μ = 2.8, standard deviation σ = 0.065) and a mean case fatality rate of 2.8% (σ = 0.64%); the model was fitted to match the number of cases of COVID-19 diagnosed in Mauritius from 18 March till 12 April 2020.

**Table 1:** Table summarizing the values of the variables used in the standard model. D = number of days needed to diagnose the first couple of persons; for the standard model, D = 28 days. N(μ, σ) represents the normal distribution with mean μ and standard deviation σ. * These variables were manually modified to make the model fit the actual data. The “first day” is defined as the first day when an infected person enters the country.

| Variable | Value |
| --- | --- |
| Population size | 1,200,000 people |
| Rate of entry of infected people* | 1 person per day for D* days; then 0 infected person per day enters the country |
| Sensitivity of point of entry screening | 64% |
| Probability a person will transmit the infection* | $\sim N(0.03, 0.04)$; lower limit = 0.01; upper limit = 1 |
| Number of contacts per person per day in the community without contact reduction | $\sim N(13, 9)$; lower limit = 0 contacts |
| Contact reduction during quarantine | Reduced by 90% |
| Contact reduction during isolation | Reduced by 95% |
| Contact reduction when universal contact reduction measures are taken* | 20% reduction for 7 days after D days; then 80% reduction; essential workers have 50% more contacts when such measures are being undertaken |
| % of population that are essential workers | 1.0% |
| Sensitivity of test for SARS-CoV-2 | 75%. Test results are available on the same day. |
| % of infected patients who remain susceptible to the virus | 14% |
| % of contacts that are successfully traced* | 25% for 7 days after D days; then 70% |
| Carrier state duration after onset of symptoms | $\sim N(8, 5)$; lower limit = 4 days. I.e. time during which test remains positive and patient can transmit infection. |
| Incubation period | $\sim N(5, 5)$; lower limit = 2 days; 0% risk of transmitting the disease in the first 2 days |
| % of tests that can become positive on a given day (positivity rate) | $\frac{25}{\sqrt[3]{P}} + \frac{20,000 \cdot A'}{P^{1.5}} - \frac{700,000,000 \cdot T_{max}}{P^2}$<br>P = population size of the country. A' = number of cases diagnosed on the previous day. T = maximum number of tests that can be done on that day.<br>Lower limit of 0.5%. Upper limit of 50%. |
| Maximum number of tests that can be done on a day | 50 tests per day for 7 days after D days; then, 100 tests daily for another 7 days; then, 200 tests daily for another 7 days; then, 400 tests daily for another 7 days; then, 500 tests each day |
| Infection fatality rate* | 1.2% |
| % of people who are asymptomatic | 5.0% |
| % of people who are mildly symptomatic | 75% |
| % of people who are moderately symptomatic | 13% |
| **% of people who are severely ill** | 6.3% |
| **% of asymptomatic people who visit the hospital** | 0% |
| **% of mildly symptomatic people who visit the hospital** | 10% |
| **% of moderately symptomatic people who visit the hospital** | 80% |
| **% of severely symptomatic people who visit the hospital** | 90% |
| **% of dying patients who visit the hospital** | 100% |
| **% of infected people who get tested for SARS-CoV-2 after visiting the hospital** | 50% for 7 days after D days; 90% afterwards |
| **% of quarantined people who get tested for SARS-CoV-2** | 100% |
| **Duration of treatment** | $\sim N(12, 2)$; lower limit of 8 days; treatment reduces the duration of carrier state by 50% if the patient is still a carrier at discharge |
| **Duration of quarantine** | 14 days |
| **Number of days needed to trace contacts** | 3 days |

**Figure 1:**
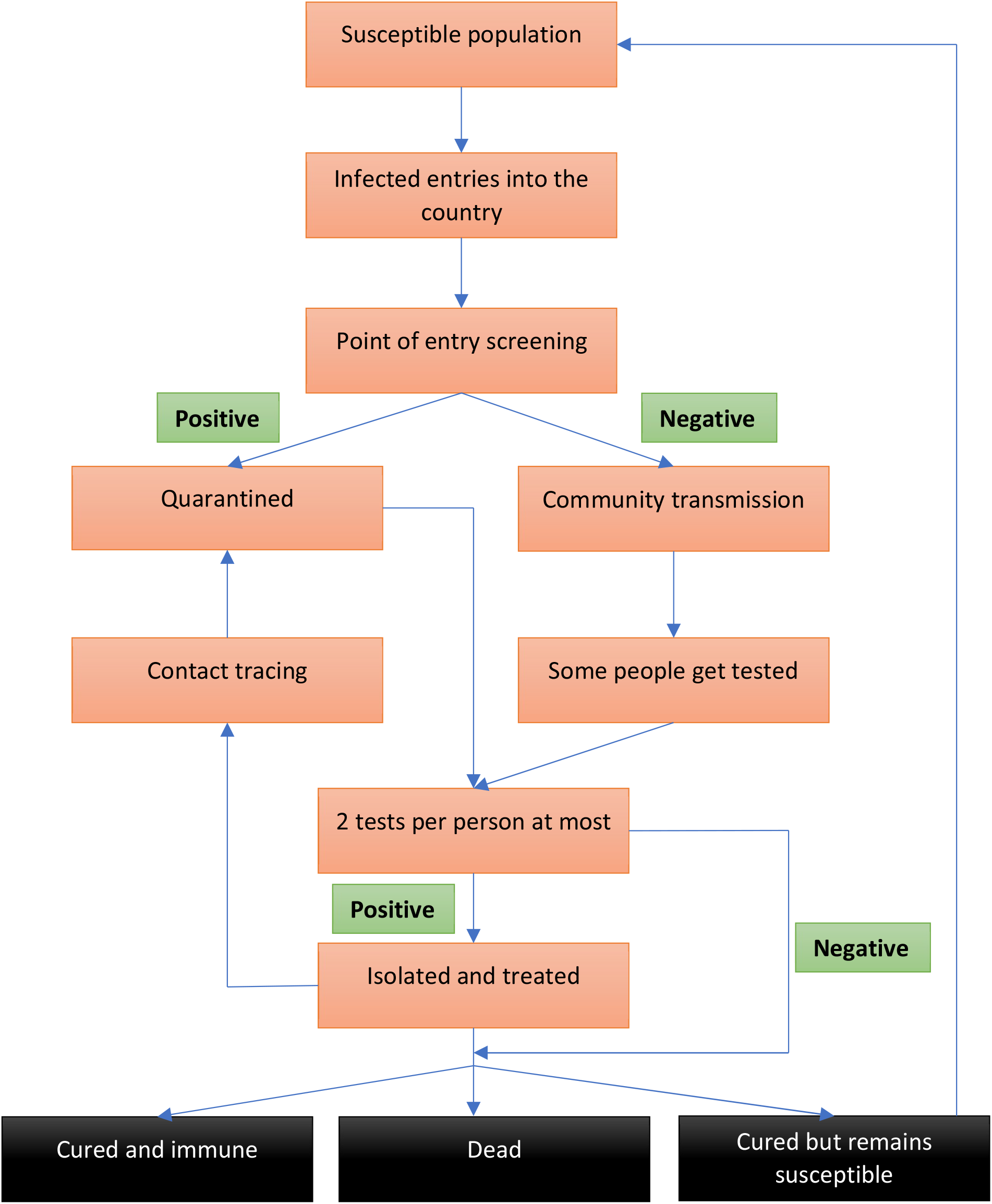
A simple schema to illustrate the model utilized in this article.

The model was run 100 times for 99 days to obtain the mean, standard deviation, and range of several variables. Multiple sensitivity analyses were conducted to ensure the results were robust. The software utilized were Excel 2003 (build 12624.20466 Click-to-Run), Java version 8 (update 181; build 1.8.0_181-b13) run on Eclipse Mars (4.5.0) and R version 3.6.1. The values of various parameters are provided in the text to within 2 significant figures.

## Results

### How well does the model resemble actual data?

Figures 2 and 3 illustrate that the model is able to simulate the actual number of diagnoses that occurred in Mauritius well.

**Figure 2:**
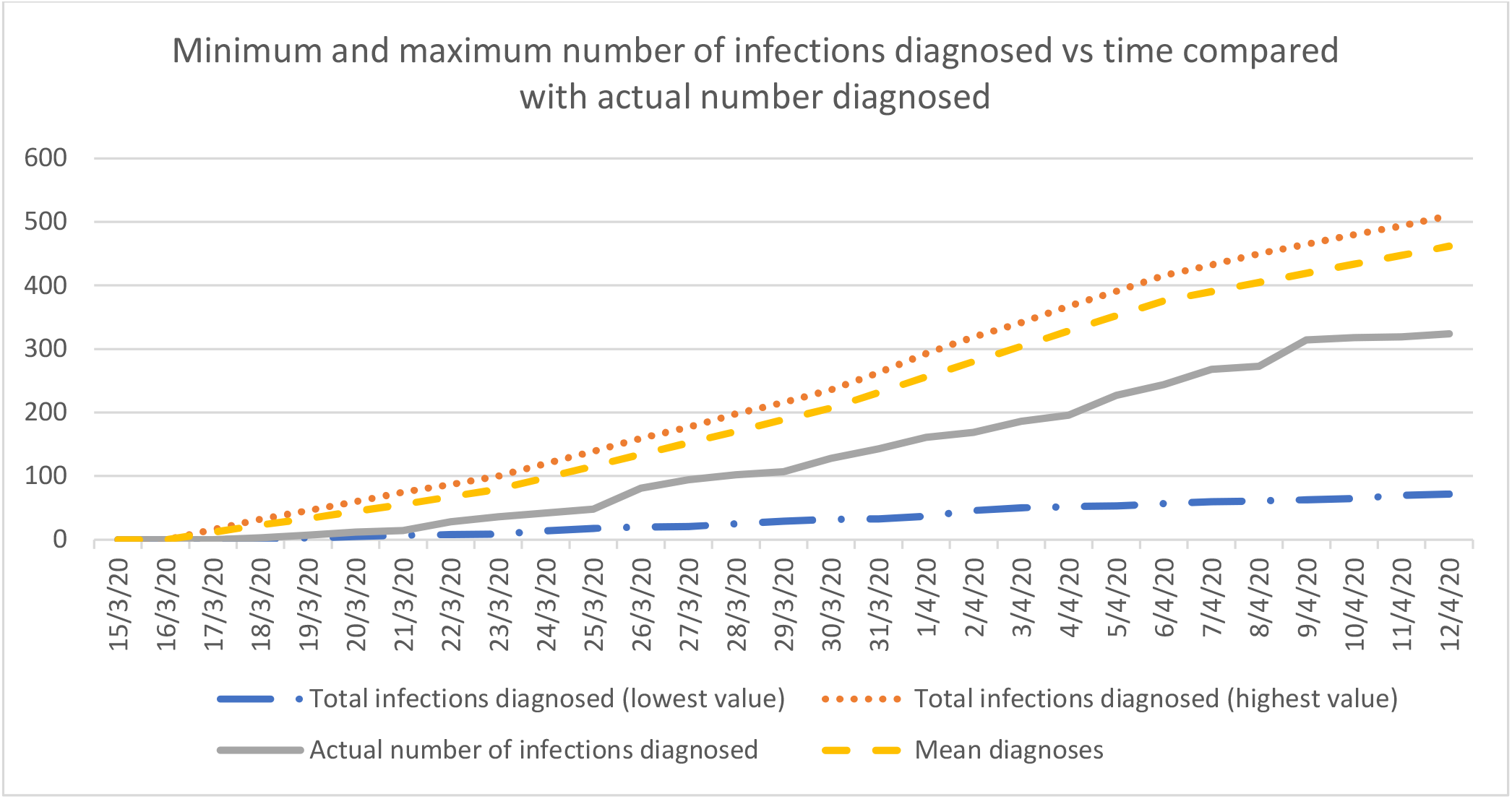
Cumulative number of cases diagnosed in the country as predicted by the standard model

**Figure 3:**
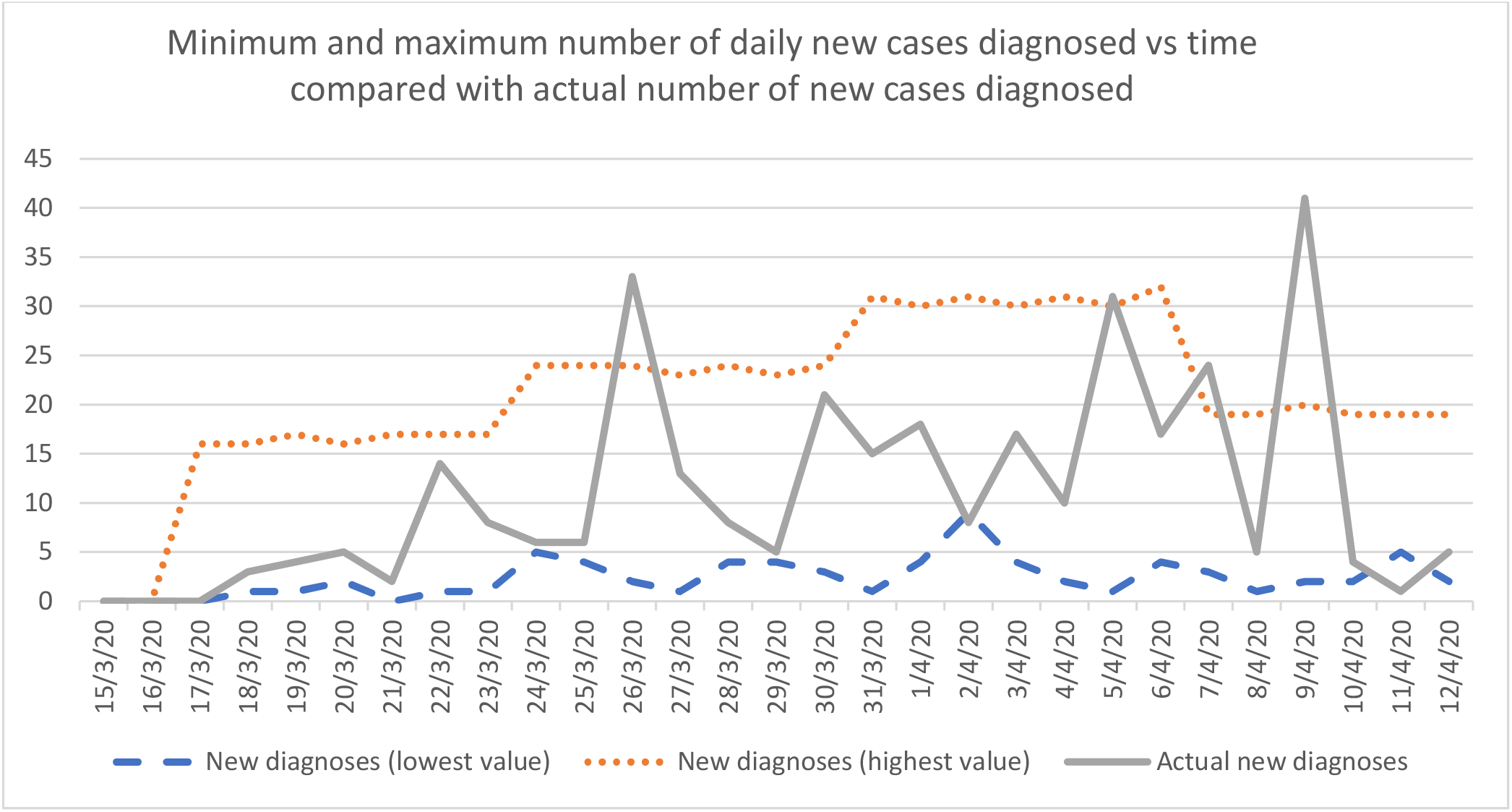
Number of daily new cases diagnosed in the country as predicted by the standard model

However, for the model to fit reality well, infections had to have started in the country 3 to 4 weeks prior to the first case being diagnosed (figure 4). This is not entirely surprising since deaths occurred soon after the first case was diagnosed, but a period of about 18 days will lapse from the time of exposure to the time of death since, based on data from China, the time from admission to death is 13 days and the time for patients to get diagnosed and admitted is slightly more than the incubation period of 5 days (2). Hence, transmission had to occur many days before the first death. Sporadic cases may have occurred even earlier without causing significant community transmission, a situation that appears to have occurred in the USA and in France (3, 4).

**Figure 4:**
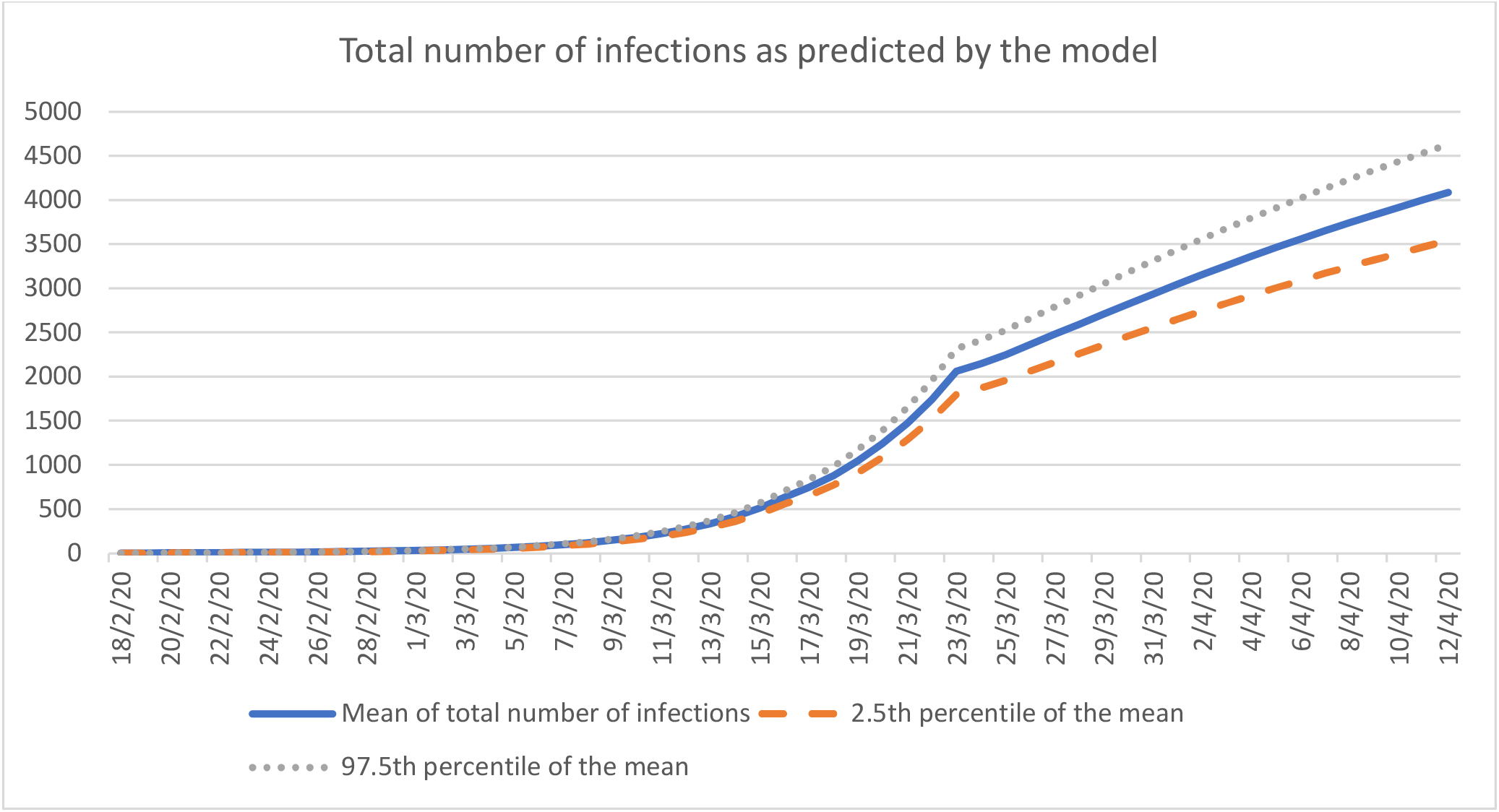
Mean of the total number of infections in the country as predicted by the model

The model predicts that the actual number of infections in the country was about 9 times higher than the number diagnosed (μ = 8.6, σ = 7.1). Moreover, the infection fatality rate was 2.3 times less than the measured case fatality rate of 2.8%.

The effects of individual public health strategies were determined from the model by examining different scenarios.

### Can testing symptomatic patients extensively stop the epidemic?

Effects on the progression of the epidemic can be measured using the effective reproductive number (R_e_). To make the model useful in practical terms across several countries, a diagnostic rate D was defined as follows:

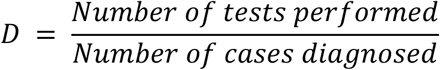

Hence, if a country diagnosed 20 new cases yesterday and performs 100 tests today, D = 5 tests per case. In the model, the least value of D was 1/10,000^th^ the population size.

Even if the test used is 100% sensitive (S = 100%) and 100% of symptomatic patients get tested (F = fraction tested = 100%), R_e_ decreases by a maximum of only 20%, a drop that occurs when D is more than 250 tests per case. For instance, when the USA had 1.5 million cases of Coronavirus Disease 2019 (COVID-19), SARS-CoV-2 patients were already identified for about 116 days in the country; for maximum effect, about 3 million tests should have been carried out per day during that period 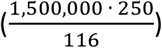, a value similar to that found in another study (5).

However, such efforts would still be insufficient to halt the epidemic since many infections are transmitted via asymptomatic or pre-symptomatic carriers i.e. R_e_ would remain above 1.

### Can mass screening of the population stop the epidemic?

In this scenario, the whole population, including asymptomatic patients, gets tested. Because mass testing will drop the positivity rate significantly, the lower limit of this rate was decreased from 0.5% to 0.25% in the model.

In the first instance, assume everyone will get tested only once while asymptomatic. Even when S and F are 100%, R_e_ drops by at most 30% when D is greater than 500 tests per diagnosis. The reason this strategy appears ineffective is (a) a large proportion of the population cannot get tested rapidly enough, meaning that some infected people will be detected later on, after they have transmitted the disease, and (b) people who have tested negative previously and are now infected, will no longer be tested, unless and until they develop symptoms i.e. they are able to propagate the disease during the pre-symptomatic phase.

We may consider another strategy: asymptomatic individuals can be tested at regular intervals for any number of times. Assuming S and F are once again 100%, R_e_ can be decreased by more than 80% when D is more than 900 tests per diagnosis; this will give a value of R_e_ of less than 1, effectively stopping the epidemic. However, assuming a more realistic value of F = 40% and S = 75%, R_e_ declines by less than 20%.

In other words, mass screening only works if a large fraction of the population can get tested at regular intervals with a highly sensitive test.

### What is the effect of contact tracing on the epidemic?

In this scenario, the test sensitivity (S) is assumed to be 75%. Symptomatic people are tested at the same frequency as stated in the standard model (i.e. mean F = 22%). Iterative backward contact tracing is applied in the model and contacts are traced within 72 hours; this simulation traces all individuals who were in contact with the patient for the duration of an incubation period. However, note that the World Health Organization (WHO) recommends tracing people who were in contact with patients for only 48 hours before symptom onset (6).

Even when 100% of contacts are traced, R_e_ decreases by at most 15% when D is larger than 250 tests per case. This can be explained by (a) the test misses several cases due to its low sensitivity, (b) insufficient tests are performed among symptomatic cases and (c) it takes several days to complete contact tracing. Of note, forward with backward tracing does not improve the results significantly.

However, if S and F are 100%, and all contacts are traced within 1 day (instead of 3 days), R_e_ can drop by about 50% (μ = 55%, σ = 15%) if tracing starts 4 weeks after the outbreak (and D > 250 tests / case); moreover, if tracing starts within 2 weeks of the outbreak, R_e_ decreases by 80% (μ = 80%, σ = 13%), which is enough to stop the epidemy.

Put in simple terms, test and trace works if the test is highly sensitive, when a large number of tests are performed, when most contacts can be traced and when it is started soon after the outbreak is identified.

### How much effective contact reduction is necessary to reduce R_*e*_ *to below 1?*

A 70% reduction in contacts has a 50% chance of reducing R_e_ to less than 1 provided the measures are taken within 14 days of the start of the outbreak and the entire population is 100% compliant. R_e_ stays low independent of the number of tests conducted, the sensitivity of the test, the fraction of people tested and the extent of contact tracing. This is similar to what is expected from herd immunity 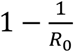. If universal contact reduction measures are implemented 28 days after the start of the outbreak, then, an 80% reduction in contacts is required, similar to what has been reported in another paper (7), to control the epidemic in less than 2 months. If compliance is not 100%, even more reduction becomes necessary.

### For how long should strict contact reduction be imposed?

It may not be realistic in most situations to severely limit contacts for prolonged periods of time e.g. via confinement. Assuming that no newly infected people enter the country, if everything returns back to normal (i.e. there is no contact reduction) 3 days after registering no new diagnoses, there is virtually a 100% chance of a new diagnosis within 2 days (μ = 2.2, σ = 0.97) i.e. a second wave is guaranteed to occur. Returning to normal 7 days after having no cases leads to a 76% chance of getting a new case within 5 days (μ = 4.9, σ = 4.2) while waiting for 14 days reduces the risk of recurrence to 28%. When the epidemy does recur in the latter scenario, it does so within 5 days also (μ = 5.4, σ = 5.5).

By estimating the daily R_e_ using various techniques (8, 9), a phased weakening of measures can be implemented; for instance when the R_e_ is consistently below 2, only a 50% reduction in contacts is necessary 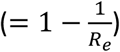. This can help to gradually ease restrictions without letting the outbreak go out of control.

The chance of a second wave is non-negligible, a point that has been demonstrated by many other models in the literature. If lockdown is the primary strategy that is used by a country to stop the epidemy, it should remain in place for more than 14 days after no cases have been diagnosed, if practically and economically feasible.

### How does point of entry screening affect the start of the epidemic?

Assuming 1 infected person enters the country per day and the sensitivity of point of entry or exit (POE) screening is 64%, there is almost a 100% that an epidemy will break out within 10 days (μ = 9.7, σ = 2.8). Even if the POE screening is 100% sensitive, outbreaks will still occur, but this will take longer to occur (μ = 20 days, σ = 10). No screening leads to an outbreak within 6 days.

POE can delay the start of outbreaks by about 2 to 3 mean incubation periods, but it does not stop outbreaks from happening, a point that has been made by other authors (10). Questionnaires and thermometers are usually inadequate (11) and for practical reasons, results of tests should be rapidly available at entry points.

### What is the effect of travel restrictions?

By reducing the entry of infected passengers to 1 person with SARS-CoV-2 entering the country every 100 days, the probability of an outbreak occurring is reduced to only 10% (over 3 months) if the sensitivity of POE screening is 100%, and to 48% if the sensitivity is 50%. However, if an epidemy does break out, it is likely to do so within the first 1 to 3 weeks.

Once again, travel restrictions serve to delay the onset of the epidemic, not to stop it entirely. Such restrictions work best when cases are identified and isolated promptly, thus preventing transmission to the community. Thompson found a risk of 41% of sustained transmission in the community from a single imported case but this risk can be reduced to 1.2% when surveillance is substantially improved (12).

### What would be the utility of population surveillance?

Suppose that a country decides to test all of its Severe Acute Respiratory Infection cases (SARI) for SARS-CoV-2 before an epidemy starts without regard to epidemiological criteria (i.e. even when the patients do not have a relevant travel or contact history). As soon as it detects its first community-acquired case, it will start all the measures as stated in the standard model i.e. travel restrictions, contact tracing, quarantine, and contact reduction.

Compared to the standard model, this strategy detects the first case sooner (at day 2 instead of day 28). The first community transmission is observed at 15 days and subsequently, the mean total number of infections at 28 days in the country is about 6 times less.

Population surveillance serves as an early warning alert and response system to allow the country to take actions quickly and to prevent a surge from happening. Countries that use mostly epidemiologic surveillance (i.e. test only travelers from high risk regions) instead of syndromic surveillance will fail to detect cases in a timely manner mostly because (a) some travelers may hide their symptoms and therefore not get tested, (b) tests are imperfect and will miss some cases among travelers and (c) pre-symptomatic transmission can happen before the traveler is diagnosed and isolated.

### What if contact reduction is not followed by a portion of the population?

Underprivileged people may not be able to remain confined for a long period of time. In this part of the model, 10% of the population continues to have twice more contacts than the remainder of the country despite strict contact reduction measures being implemented. Under such circumstances, the probability of the epidemy ‘ending’ within 99 days (i.e. reaching consecutive 3 to 14 days without any cases) decreases by 3 to 10 times when compared to the standard model.

This implies that compliance to universal contact reduction measures is critical for success. The less compliant part of the population can act as reservoirs and re-introduce infections which will prevent the outbreak from ending.

## Discussion

In a disease that can spread effectively during the pre-symptomatic phase, large-scale testing of symptomatic patients will not stop the epidemy. However, testing remains important to identify at-risk individuals who may benefit from treatment.

On the other hand, universal screening of asymptomatic individuals can help halt the progress of the outbreak, but it may not be cost-effective since a large proportion of the population must be regularly tested, and it requires a test with sensitivity greater than 90%. This may not be practical on its own.

While early, backward, iterative contact tracing together with comprehensive testing of symptomatic cases and quarantining of contacts can have significant effects on the incidence curve, this is still insufficient to decrease the effective reproductive number below 1 especially if the R_e_ is more than 5. When examining a more realistic situation in which 80% of contacts are traced, the sensitivity of the test is 90%, 90% of symptomatic patients get tested and test and trace starts 2 weeks after the first infected person enters the country, this strategy will work only if R_e_ is less than 2. Ideally, test and trace should be combined with other strategies in order to be successful, a finding that is confirmed by other authors (13).

Once the exponential phase of the outbreak starts, in virtually all scenarios where R_e_ is above 2 and detection of community transmission is delayed by more than 2 weeks, universal contact reduction strategies are required to stop a surge. Since lockdowns are not economically viable, it is imperative for countries to invest in what are sometimes viewed as inconvenient measures like social distancing and universal masking. The objective is to reduce the risk of transmission per contact or the number of contacts per person to such a low value that the epidemy will halt. For instance, preliminary evidence indicate that physical distancing or masking may reduce transmission risk by 60% to more than 80% (14), which suggest these measures may work well if R_e_ is less than 3 to 5.

Such contact reduction techniques should be continued for as long as possible until after the outbreak dies out, preferably 28 days after the last case was diagnosed since this reduces the risk of a second wave to less than 10%. However, it may not be economically feasible to maintain universal contact reduction measures for a prolonged period and relaxation of these measures can be entertained as long as the estimated daily R_e_ is gradually decreasing; a strong test and trace system will ensure that R_e_ does not rise again.

In a situation where most parts of the world are still being challenged by COVID-19, re-importation of the disease is highly likely once the lockdown is eased; many models suggest that flattening the curve using lockdowns can delay the peak of the epidemy but the total number of infections remain the same or in worst case scenarios, it may even increase if lockdowns are started too late (15). While lockdowns can provide precious time initially to prepare the public and the healthcare system to face the disease, this is a reasonable long-term solution only if the lockdown is relaxed when (a) herd immunity is reached e.g. via effective vaccination with minimal side effects and long-term protection or (b) the country no longer registers new cases and its borders can remain closed until most parts of the world have controlled the epidemic. If these conditions are unlikely to be met (e.g. in the case of a widespread pandemic during which R_e_ is greater than 1 in many countries and worldwide collaboration appears inadequate to bring the global R_e_ below 1 rapidly), some epidemiologists have advocated the use of controlled herd immunity which can be successful if (a) most patients become immune to the disease on recovery, (b) the vulnerable part of the population can be adequately protected from the infection and (c) the infection is not allowed to surge so as not to overwhelm the health services and also to give enough time for neutralising antibodies to form. Although controlled herd immunity was not thoroughly assessed in this model, assuming functional antibodies take 14 days to form, R_e_ should be maintained between 1.0 and 1.7 for this strategy to work – it can take several months to years to reach herd immunity thus. Sweden may have failed in using this strategy partly because it did not reduce its R_e_ sufficiently (e.g. via extensive contact tracing) and it did not protect the frail segment of its population appropriately.

To minimize the risk of a second wave from imported cases, all individuals entering the country should undergo POE screening. Alone, this is unlikely to have a substantial impact, especially since current tests have less than 90% sensitivity. Some form of travel restrictions should also be considered if the country is currently not well prepared to face a second wave. Travelers may be allowed to enter the country if they come from an area with a low prevalence of the disease.

Once the epidemy is over, an early warning system is of paramount importance. This may involve sentinel surveillance, random population screens and / or testing patients with influenza-like illness or SARI for SARS-CoV-2. Once again, it is necessary to ensure that the quality of the test is good or else too many cases will be missed. While the model emphasizes the significance of sensitivity, specificity is equally salient; doing a test with poor specificity on many patients will lead to frequent false positives that can drain resources from the healthcare system.

In many countries, the destitute and needy may not have access to masks or alcohol sanitizers. They live in crowded areas like slums and ghettos. Such conditions can encourage the transmission of SARS-CoV-2. By accounting for this segment of the population in the model, it was shown that neglecting this group of people leads to a slower decline in the epidemy – equality of access to healthcare is crucial.

Figures 5 and 6 illustrate some key steps that should be taken by a country when facing an outbreak from primarily a respiratory illness like COVID-19. These can help countries develop appropriate policies. Some fundamental points must be emphasized:

**Figure 5:**
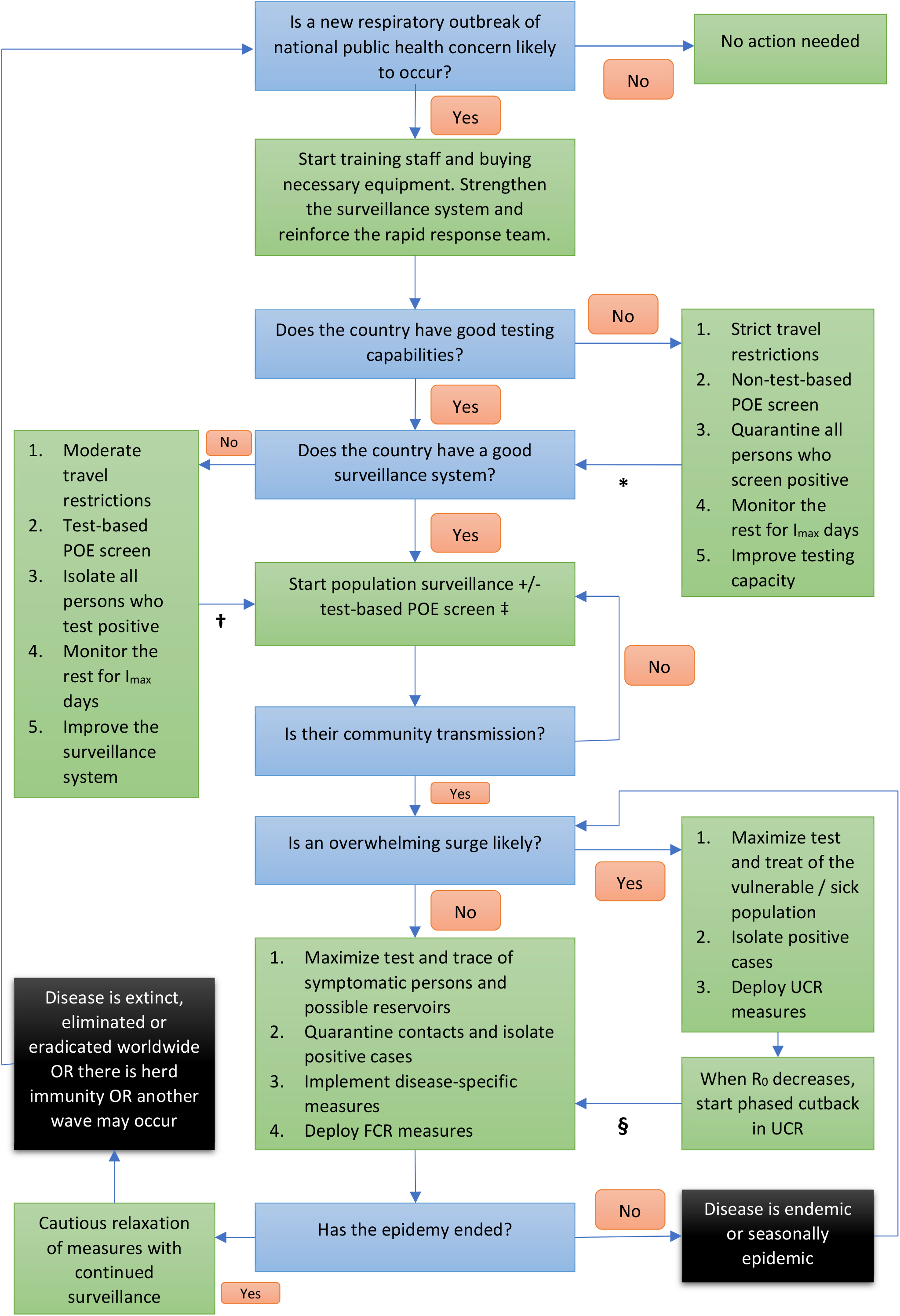
Measures that should be considered when an outbreak might occur in a country. POE = point of entry and/or exit; I_max_ is the maximum duration of the incubation period or its 99^th^ percentile, whichever is more practical (I_max_ = 14 days for SARS-CoV-2); R_0_ is the basic reproductive number; UCR = universal contact reduction; FCR = focused contact reduction. * = go to the next box when testing capability is adequate or after I_max_ days has passed after time zero (in the latter situation, keep strict travel restrictions until better testing capability is available); † = go to the next box when population surveillance is adequate or after I_max_ days has passed after time zero (in the latter situation, keep moderate travel restrictions until better population surveillance is carried out); ‡ = carry out test-based POE screening if the anticipated surge of cases is likely to overwhelm the healthcare system; § = go to the next box when R_0_ is persistently and significantly less than 2. Time zero is the time when the country is most likely to see its first infected person and in case this is too difficult to ascertain, it may be taken to be the time when the WHO declares a Public Health Emergency of International Concern. To the extent possible, use a combination of strategies to have maximum effect. See table 2 for details about the terms used in this figure.

**Figure 6:**
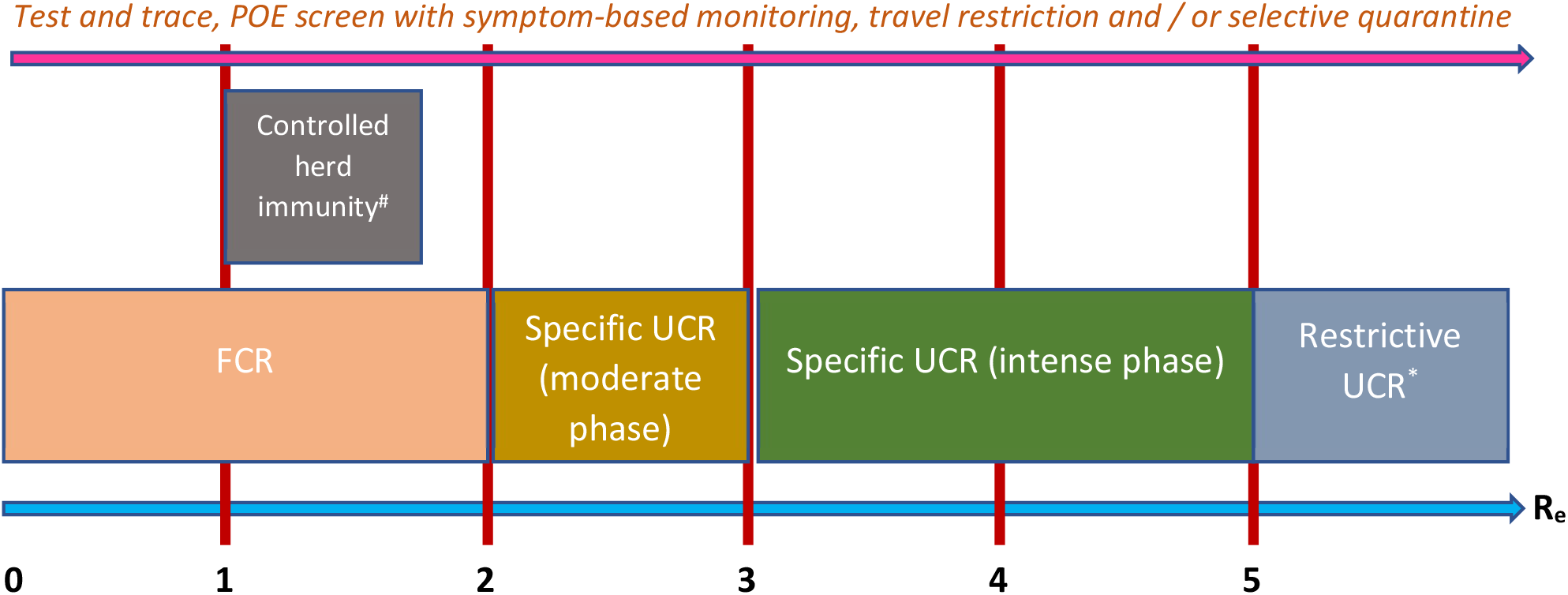
Summary of strategies that may be used during the exponential phase of a COVID-19 epidemic. Factors other than just the effective reproductive number should be considered before deciding which strategy to implement. POE – point of entry or exit; FCR – focused contact reduction; UCR – universal contact reduction. # - to implement if there is evidence of effective, long-term immunity after the infection, the vulnerable part of the population can be properly protected and R_e_ can be kept under 1.7. * - to implement if benefits outweigh harms. Intense phase - ≥ 80% of the population should decrease their number of contacts by ≥ 80%; moderate phase - ≥ 80% of the population should decrease their number of contacts by ≥ 60%. Test and trace work best when applied early and when R_e_ is less than 2. See main text regarding how to optimally use POE screen and travel restrictions. For infections that have milder health impact, the graph should move to the right i.e. use more stringent measures when R_e_ is higher. See table 2 for details about the terms used in this figure.

– Deciding which outbreaks are concerning enough to trigger potentially economically ruinous interventions is not easy. Most countries rely on the WHO to declare a Public Health Emergency of International Concern, but the algorithm that the WHO uses should be improved (16).
– Equally problematic is determining when an outbreak is getting out of control. An overwhelmed healthcare system is associated with a higher mortality. Swift and decisive actions must be adopted by the country under such circumstances to mitigate the damage. A short doubling time of the number of cases together with a sharp drop in the number of beds available in hospitals should be cause for concern.

Had Mauritius followed the steps in figure 5 rigorously, the country would have seen 6 times less cases of COVID-19 and virtually zero deaths.

**Table 2:** This table describes some of the terms used in figures 5 and 6. Numbers that are provided here should be used as a guidance only. If some of the criteria cannot be fully met, then compensation should be sought in some other way e.g. if the sensitivity of the test is much less than 90%, then much more than 80% of contacts should be traced.

| Terms | Comments |
| --- | --- |
| **Respiratory outbreak** | The model simulates outbreaks due to respiratory illnesses. The algorithm illustrated in figure 5 should not be used for other infections that are transmitted by the fecal-oral or vector-borne routes. These diseases require different measures to be employed. |
| **Outbreak of national public health concern** | Typically, this represents a new disease that is not already endemic in the country, that can spread rapidly in the community and that can have a major economic and health impact on the population. The country should raise the alert if it believes it is vulnerable to such an infection. |
| **Good testing capability** | This implies that the country has in sufficient amount a test that is reasonably sensitive and specific (typically more than 90% sensitive and more than 90% specific). The number of tests needed varies depending on the reproductive number, transmission duration, testing strategy used, health impact of the disease and countermeasures in place. If the disease has low mortality and causes mild symptoms, less people need to get tested. As a rough guide, initially, countries may prepare to test about 20% of its population annually. Testing needs should be evaluated regularly. |
| **Travel restriction** | Strict travel restriction means less than 1 infected person enters the country every 100 days while moderate travel restriction allows 1 infected person to enter the country every 10 days. The benefits of restriction must be balanced with the economic harm it can cause. Travel restrictions only delay entry of disease into the country by about 2 to 4 mean incubation periods for every 10 days that an infected person is prevented from entering the country. However, this strategy gives time for the country to prepare itself and both strict and moderate restrictions reduce the burden of disease early on by about 4 times. If a country has C known active carriers and a population P', then to allow 1 carrier to enter every N days, the destination country should accept $\frac{P'}{N \cdot C \cdot k}$ persons daily; C·k represent the total number of carriers (including the undiagnosed ones) and when this number is unclear, k may be estimated to be 10. Use of travel bubbles (travel bridges or Corona corridors) may help. For example, France had 54,818 active cases on 1 June 2020 – if N = 10, almost 100 passengers could be allowed to enter the country from France every week (after point of entry or exit screen). |
| **Non-test-based point of entry or exit screen** | Point of entry and / or exit screen only delays the start of an outbreak and is less effective than travel restrictions. Non-test-based screening focuses on patient symptoms, temperature, and country of origin as well as countries recently visited and other epidemiologic criteria. All travelers that come from countries where community transmission is suspected or confirmed should be screened. |
| **Test-based point of entry or exit screen** | Test-based screens utilize point-of-care lab tests that should preferably have more than 90% sensitivity and more than 90% specificity. All travelers that come from countries where community transmission is suspected or confirmed should get tested. Due to the expense involved, use this type of screening only if the healthcare system is unable to respond to the upcoming surge. Test-based screens can reduce the number of infections early during the epidemic by about 3 times. |
| **Quarantine** | For practical reasons, selective quarantine is usually employed: people who are recently in contact with confirmed or suspected cases are quarantined (self / home or institutional). Symptomatic persons are isolated (self / home, institutional or healthcare-based). If asymptomatic transmission is prevalent and the screening tool has poor sensitivity, general quarantine may be considered within the limits of |
|  | acceptability i.e. all travelers from certain highly afflicted countries may be quarantined. Institutionalized quarantine is preferred by some authorities, but this must be balanced with the cost involved and the risk of enhanced transmission inside an enclosed setting. |
| **Monitoring of travelers** | Travelers should get re-tested if they develop symptoms within the incubation period. Monitoring can be active (whereby a healthcare worker checks on the person at a certain frequency) or passive (= self-monitoring). If resources permit, active monitoring is preferred. |
| **Good population surveillance system** | Active (= sentinel) surveillance should be favored over passive surveillance. Usually, syndromic surveillance is carried out; however, sewage testing and pool testing with polymerase chain reaction can be useful. In this context, a good system should detect the start and the extent of community transmission as early as possible. In situations where asymptomatic, pre-symptomatic or mildly symptomatic patients can transmit the infection effectively, a large number of people must be tested irrespective of travel or contact history e.g. > 90% of patients with symptoms suggestive of the infection should be tested even if epidemiologic criteria are not met since community transmission will remain otherwise undetectable. |
| **Overwhelming surge** | This term suggests that the healthcare facilities may no longer be able to provide the services needed to people who fall sick. The country should create models to estimate under what conditions this is likely to occur. Accurate data must be collected daily over the period of 1 to 2 (or more) mean incubation periods - as a rough estimate, if $R_e$ is estimated to be greater than 3 or the doubling time is less than 7 days as well as the number of free beds in the hospitals has dropped by more than 30% compared to the usual number of free beds available during that time of the year, an overwhelming surge may be imminent in that area. Under such circumstances, countries should consider expanding the healthcare capacity to the maximum possible before applying restrictive universal contact reduction. |
| **Universal contact reduction** | Such measures can be divided into *restrictive* ones like lockdown (intermittent or continuous; preventing people from getting out of their homes), confinement (preventing people from getting out of an area) and curfew (preventing people from getting out at certain times of the day), and *specific* ones like wearing of masks among asymptomatic people or when in crowds, frequent hand sanitizing, unrestricted gloving, regulating crowd size and universal social distancing. Restrictive UCR may be considered when $R_e$ is too high (e.g. > 5) to control the outbreak using other strategies (for an infection that can cause an unacceptable number of deaths or harm) or during the early phase of the outbreak when transmission dynamics and epidemiological characteristics are being elucidated. |
| **Focused contact reduction** | Such measures include the closure of high-risk locations e.g. schools, universities or some workplaces, shielding or cocooning the vulnerable segment of the population (e.g. by wearing masks around them and social distancing from them), not coming to work when having respiratory symptoms (i.e. using paid sick leaves), wearing masks when symptomatic, working from home or tele-work, distancing from people who cough, and applying good infection prevention and control within high-risk settings like healthcare facilities. |
| **Test and trace** | When performed properly, more than 80% of patients who present with symptoms suggestive of the disease should be tested, more than 250 tests should be done per diagnosis (while community transmission is occurring) and more than 80% of contacts should be traced and quarantined within 72 hours. If test and trace is started within |
| | 1 mean incubation period after the beginning of community transmission, the chances of controlling the outbreak are particularly good. If this strategy is delayed, the chances of success diminish considerably but $R_e$ may still be reduced by about 50% instead of 80%. This strategy fails when too few tests are performed. When the incubation period is relatively short (e.g. < 1 month), testing of random asymptomatic people is not practical since too many people need to get tested in too short a period of time in order to have any effect on the outbreak. |
| **Reservoirs** | Reservoirs can include animals or vectors. Moreover, the poor and the marginalized people in the society may not get tested nor treated appropriately; they can act as reservoirs and prevent an outbreak from ending. |
| **Phased cutback of measures** | Due to their impact on the economy, restrictive universal contact reduction measures should be eased as soon as possible. When $R_e$ is around 2 consistently, contacts can be reduced by only 50%; specific universal contact reduction may be used for this purpose instead of restrictive ones; test and trace as well as focused contact reduction should be emphasized when $R_e$ is less than 2 for 1 to 2 $I_{\max}$ (the maximum incubation period). The use of social bubbles may help. |
| **Disease-specific measures** | These measures can include immunization with a reasonably effective vaccine (and with minimal side effects), early test and treat (to reduce the viral load which can then decrease the transmission risk), pre-exposure prophylaxis or post-exposure prophylaxis. |
| **End of epidemic** | The outbreak is considered to have ended if there are no active cases in the country for the duration of 2 $I_{\max}$ days. To ensure this is the case, adequate testing must be carried out. |

Even though all models are ultimately wrong and all countries are facing their own challenges, I hope that the model that was utilized in this article has highlighted the importance of good population surveillance and the need to keep a low number of contacts for an extended duration of time. Furthermore, it is a priority to look for an affordable diagnostic test for SARS-CoV-2 that is more than 90% sensitive and specific.

## Data Availability

No data are available.

## Appendix: Section A Justifications for the values of the variables used in the model

**Appendix: Section A – Justifications for the values of the variables used in the model**
| **Variable** | **Value** |
| --- | --- |
| **Population size** | The population of Mauritius in 2019 was 1,269,668 (1) |
| **Rate of entry of infected people** | The borders of Mauritius were closed soon after COVID-19 cases were identified in the country |
| **Sensitivity of point of entry screening** | Quilty et al. describes a 64% sensitivity of screening for SARS-CoV-2 at the airport (2) |
| **Probability a person will transmit the infection** | This was set so that the model gives a value of $R_0$ equal to about 3, which is a number frequently quoted in the literature (3). This $R_0$ is calculated from the infected population at 28 days. The theoretical $R_0$ calculated from transmission data is otherwise 4.29 in the model. |
| **Number of contacts per person per day in the community without contact reduction** | Mossong et al. suggests that people have a mean number of 13 contacts per day in the community (4) |
| **Contact reduction during quarantine** | Assumption |
| **Contact reduction during isolation** | Assumption |
| **Contact reduction when generalized public health measures are taken** | This was set so that the mean number of infections that was diagnosed 1 month after the first case was identified would be from 300 to 500. |
| **% of population that are essential workers** | Assumption |
| **Sensitivity of test for SARS-CoV-2** | Tests based on polymerase chain reaction (PCR) are used to diagnose patients in Mauritius. The sensitivity is unknown. While in-vitro sensitivity can be higher than 95%, in-vivo sensitivity is much lower at 56% to 83% (5). Throat swabs may have a lower sensitivity of 32% compared to nasopharyngeal swabs (6). In Mauritius, from March to April, most patients were getting tested with throat swabs with each person expected to get swabbed twice per test. Assuming a sensitivity of 50%, a set of 2 such swabs can be said to have a sensitivity of 75%. |
| **% of infected patients who remain susceptible to the virus** | Song Tie, vice director of the local disease control center in southern China's Guangdong province announced on 26 Feb 2020 that China has noted a re-infection rate of 14% (7) |
| **% of contacts that are successfully traced** | This was set so that the model accurately predicts the number of diagnosed cases during the first month after the first infected person is identified |
| **Carrier state duration after onset of symptoms** | According to the technical report from the European Center for Disease Prevention and Control (ECDC), the viral load of SARS-CoV-2 persists for 8 to 11 days after symptom onset (8) |
| **Incubation period** | The incubation period has been estimated to be from 4.5 days to 5.8 days, but some patients may develop symptoms after 14 days (9) |
| **% of tests that can become positive on a given day (positivity rate)** | After analyzing statistics on SARS-CoV-2 from the USA and from Mauritius, an equation was empirically and manually created to relate the positivity rate with the size of the population, the number of cases diagnosed and the number of tests performed. This equation may not apply to other countries. |
| **Maximum number of tests that can be done on a day** | This was based on the actual number of tests that could be carried out in Mauritius at the time. |
| **Infection fatality rate** | This was set to give a case fatality rate (number of identified deaths from SARS-CoV-2 over total number of cases diagnosed) of about 3% which is the mortality rate observed in Mauritius at the time. |
| **% of people who are asymptomatic** | The ECDC suggests that the true asymptomatic rate may be from 1% to 3% (10). This excludes pre-symptomatic or mildly symptomatic patients. Other estimates can go up to 50% although it is likely that these values include patients in the pre-symptomatic phase (11). |
| **% of people who are mildly symptomatic** | About 80% of patients have mild symptoms according to Worldometer (12) |
| **% of people who are moderately symptomatic** | 2/3 of the people who remain (i.e. not asymptomatic nor mildly symptomatic nor dying) were assumed to have moderate symptoms in the model |
| **% of people who are severely ill** | The rest of infected patients have severe symptoms (but do not die) |
| **% of asymptomatic people who visit the hospital** | Asymptomatic patients have no reason to visit the hospital |
| **% of mildly symptomatic people who visit the hospital** | Patients with few symptoms rarely visit the hospital. |
| **% of moderately symptomatic people who visit the hospital** | Most of these patients will visit the hospital |
| **% of severely symptomatic people who visit the hospital** | Most of these patients will visit the hospital |
| **% of dying patients who visit the hospital** | All such patients will visit the hospital according to the model (although there are reports of patients dying without reaching any healthcare facilities) |
| **% of infected people who get tested for SARS-CoV-2 after visiting the hospital** | At the start of the outbreak, healthcare facilities were still getting prepared and patients were not getting tested as often as needed. Later, most patients with respiratory symptoms were getting tested. |
| **% of quarantined people who get tested for SARS-CoV-2** | All quarantined patients should get tested twice. This was not the case initially in Mauritius but eventually became the standard. |
| **Duration of treatment** | Many patients stayed in the hospital for about 14 days in Mauritius. Patients were discharged only after 2 consecutive negative PCR tests. Xiao et al. noted that it can take 17 to 24 days for the PCR to become negative after onset of symptoms (13); it takes a few days after the start of symptoms for patients to seek hospital care. Fonfria et al. suggest the length of stay in the hospital is from 13 days to 15 days (14). |
| **Duration of quarantine** | As per WHO recommendations, the duration of quarantine is 14 days. |
| **Number of days needed to trace contacts** | Assumption |

## Appendix: Section B Additional assumptions used in the model

1. The population remains static i.e. births, deaths and migration do not significantly affect the number of people in the country due to the small changes involved.
2. 0% of the population has immunity to SARS-CoV-2 at the start of the outbreak.
3. All infected travelers who enter the country are assumed to be at the start of their incubation period.
4. The transmission rate of the virus does not vary with time nor with symptoms in the model. However, in reality, it is believed that patients who have more severe symptoms can transmit the virus more effectively, while patients who are asymptomatic may be inept spreaders.
5. Risk of death of any person is the same i.e. demographic characteristics like age or co-morbidities are not considered.
6. Transmission throughout the population is taken to be homogenous in the model. Nonetheless, it is well established that networks exist within various regions of a country in which transmission may be more proficient.

## Appendix: Section C Model structure

**Figure 7:**
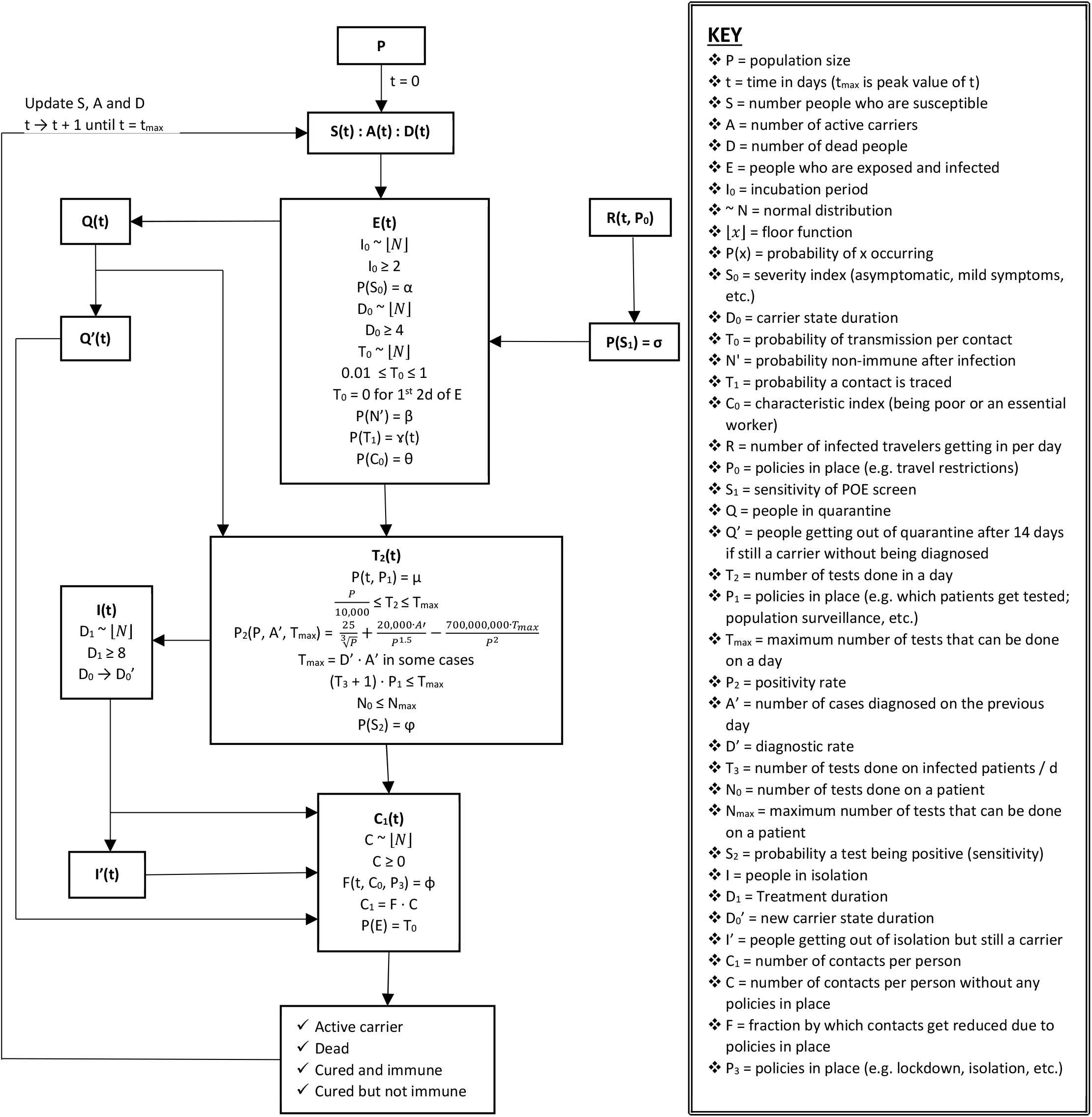
The mathematical and programmatic structure of the standard model. See the main article for values and definitions of variables. When a variable that follows a probability distribution exceeds its limits, it is approximated to the nearest bound. Q(t) includes patients who are screened positive through S_1_ or through contact tracing T_1_. I(t) includes people who are tested positive. Patients remain a carrier at least till the end of treatment (if treated). POE = point of entry or exit. Calculated reproductive number at time t = number of secondary infections from people who are no longer carriers (including dead people) at time t / total number of these people at time t.

